# Application of 4D flow MRI for exploring the anatomical and haemodynamic characteristics of pelvic veins and their correlations

**DOI:** 10.1101/2023.07.17.23292798

**Authors:** Chengli Nie, Li Tao, Jiangwei Chen, Jianghu Yang, Wen Huang

## Abstract

**Objective:** There is currently a lack of studies evaluating the anatomy and haemodynamics of the pelvic veins in clinical practice. Four-dimensional flow magnetic resonance imaging (4D flow MRI) can simultaneously obtain information on vascular anatomy and haemodynamics. The goal of this study was to use 4D flow MRI to explore the anatomical and haemodynamic characteristics of pelvic veins and their correlations in an asymptomatic population.

**Methods:** Four-dimensional flow MRI was used to measure anatomical parameters, including the pelvic vein area, common iliac vein-inferior vena cava angle, iliac vein tortuosity, and common iliac vein stenosis rate, and haemodynamic parameters, including the pelvic venous blood flow, average blood flow velocity, and external iliac vein-inferior vena cava pressure difference, in 30 asymptomatic volunteers. The correlation between iliac vein anatomy and haemodynamics was analysed.

**Results:** There were correlations between the anatomical parameters of the iliac vein and the haemodynamic parameters. A larger iliac vein area was correlated with a higher blood flow. A greater iliac vein stenosis rate was correlated with a lower blood flow, lower mean velocity of the external iliac vein, and higher external iliac vein-inferior vena cava pressure difference. A greater common iliac vein-inferior vena cava angle was associated with a lower iliac vein flow velocity and a higher external iliac vein-inferior vena cava pressure difference. Finally, a greater the iliac vein tortuosity was correlated with a lower iliac vein velocity and higher external iliac vein-inferior vena cava pressure difference. There were differences in the anatomical and haemodynamic parameters of the bilateral common iliac vein and external iliac vein. The left pelvic vein common iliac-inferior vena cava angle, iliac vein tortuosity and iliac vein stenosis rate were higher than those of the right side. The flow and average velocity of the left common iliac vein and the left external iliac vein were lower than those of the right, and the pressure of the left external iliac vein was higher than that of the right.

**Conclusion:** The anatomical structure of the pelvic vein, including the iliac vein area, common iliac vein-inferior vena cava angle, iliac vein tortuosity, and iliac vein stenosis rate, are important factors that affect haemodynamic changes in the pelvic vein. There are correlations between parameters related to the anatomical structures and haemodynamic changes of the pelvic veins. Differences in the anatomy and haemodynamics of the bilateral pelvic veins may be one reason why the left extremity is more prone to chronic venous disease (CVD)-related symptoms.

## 1. Introduction

Chronic venous disease (CVD) of the lower extremities is a common clinical syndrome whose incidence increases with age. Patients often present with symptoms such as lower extremity heaviness, lethargy and swelling and signs such as lower extremity varicose veins, lower extremity oedema, skin pigmentation, and venous ulcers. The reason for these symptoms is the development of lower extremity venous stasis caused by various factors, including obstructive factors such as pulmonary hypertension, heart failure, Budd-Chiari syndrome and iliac vein stenosis and regurgitation factors such as tricuspid regurgitation and lower extremity venous valve insufficiency. Clinical studies have found that iliac vein stenosis is an important factor leading to venous reflux disorders in the lower extremities, but it is not clear how it leads to changes in iliac vein haemodynamics. Iliac vein stenosis can be identified and treated in clinical practice, but sometimes these treatments cannot relieve the symptoms of lower extremity venous insufficiency. Therefore, it is clinically important to understand the characteristics of pelvic vein haemodynamics.

4D flow MRI can simultaneously obtain vascular anatomy and haemodynamic information and dynamically display haemodynamic changes, including blood flow velocity, blood flow shear force, and kinetic energy. It can also visualise the direction and state of blood flow in the form of flow diagrams and trace diagrams and aid in quantitatively and qualitatively analysing blood vessels^1^, which can help in investigating the causes of vascular disease and the factors affecting disease progression. In addition, by excluding contrast enhancement from 4D flow MRI analysis, its interference on blood flow is excluded, resulting in a more realistic reflection of the haemodynamic state of the body.

We used 4D flow MRI for the first time to investigate the haemodynamic characteristics of the iliac veins in young asymptomatic people. We intend to study baseline pelvic vein anatomy and haemodynamics data and provide a basis for further research on the pathogenesis of CVD of the lower extremities.

## 2. Method

### 2.1 Study subjects

This study was approved by the Research Ethics Committee of our hospital. A homemade questionnaire was distributed to young volunteers from September 2022 to March 2023 to determine whether they have symptoms related to chronic venous insufficiency of the lower extremity and to determine whether the lower extremities had signs of oedema, varicose veins, skin pigmentation and/or venous ulcers by physical examination. Subjects were recruited according to the results of the questionnaire and physical examination according to the following inclusion criteria:(1) no vascular-related symptoms (swelling, oedema, hyperpigmentation and venous ulcers) or confirmed vascular disease (ulcers varicose, deep vein thrombosis, thrombophlebitis); (2) body mass index (BMI)<28kg/m^2^; (3) no vein-related pressure or drug treatment; (4) no contraindications related to MRI scanning. Four-dimensional flow MRI examinations were performed after obtaining informed consent from the included subjects. The exclusion criteria were congenital heart valve insufficiency, anatomic abnormalities of the inferior vena cava and iliac vein, a venous thromboembolism (VTE)-related family history, and lateral branch formation. The demographic and medical records of the volunteers were collected, and their initial clinical status was assessed according to the VCSS and Villalta scores.

### 2.2 Anatomical and haemodynamic parameters

Anatomical images were obtained with an MR device (Ingenia DNA 3.0T, Philips, The Netherlands) using the Qflow_BH sequence during end-expiratory breath-hold with the patient in the supine position at standard coronal and sagittal orientations, starting 1 cm above the iliac vein bifurcation (slice thickness 8 mm). QFLOW analysis software was used to initially measure the inferior vena cava blood flow velocity Vc, and the velocity encoding (VENC) value was calculated as Vc*(1 ± 20%). Haemodynamic information from the inferior vena cava, bilateral common iliac veins and proximal external iliac veins for 1 cardiac cycle was acquired using the 4D-FLOW-PC sequence in the axial orientation. After the original sequence scan was completed, two sets of data, the modular (M) image and the phase contrast MR angiography (PCA) image, were automatically generated. The PCA image was removed, and the M image of the first group and the reconstructed phase (P) image in the right-left (RL), foot-head (FH), and anterior-posterior (AP) directions were used for postprocessing analysis.

### 2.3 Anatomical parameters: vessel area, stenosis rate, common iliac vein-inferior vena cava angle, and tortuosity

To determine the vessel area, vessel regions of interest (ROIs) were delineated at the distal end of the inferior vena cava, the proximal end of the bilateral common iliac vein (wherein the proximal end of the left common iliac artery is the distal end of the right common iliac artery crossing the left common iliac vein), and the proximal vessel area of bilateral external iliac vein.

Stenosis rate: We used the area stenosis rate of the vessel in the axial orientation to calculate the vessel stenosis rate. In the axial plane, we measured the areas where the vessel was narrowest (Smin) and widest (Smax) at the distal end of the common iliac vein, attempting to avoid the bifurcation of the common iliac vein and the site where it converges into the inferior vena cava when selecting the largest plane. Then, the vascular stenosis rate was calculated as 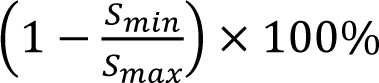.

Common iliac vein-inferior vena cava angle: Using the postprocessed coronal 4D flow MR images, the angle between the central line of the bilateral common iliac vein and the extension line of the inferior vena cava was measured as the iliac-inferior vena cava angle (Figure 1).

**Figure 1:**
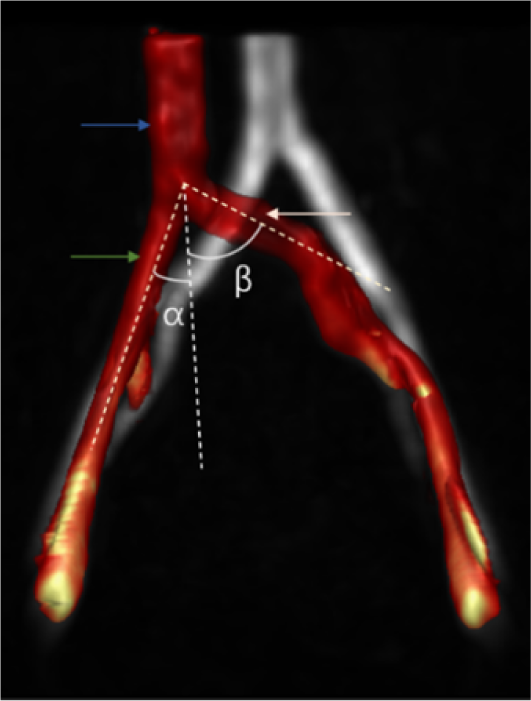
The blue arrow is the IVC, the white arrow is the LCIV, the green arrow is the RCIV. The angles α and β are the angles of the extension lines of the inferior vena cava and central line of right and left common iliac vein.

Tortuosity: The length L between the two ends of the iliac vein and the actual vessel length H were measured and used to define the tortuosity as S=H/L*100%(Figure 2).

**Figure 2:**
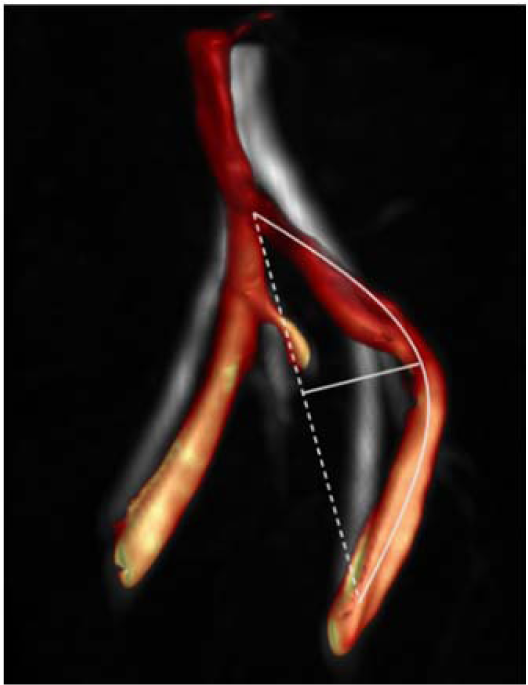
Length of iliac vein center line is L and maximum chord height is H

### 2.4 Haemodynamic parameters: flow, velocity, pressure difference

After the vascular ROIs were delineated on cvi42 from Circle Cardiovascular Imaging (Version 5.14, Calgary, Canada), the following parameters were calculated; blood flow and average flow velocity at the distal end of the inferior vena cava, the proximal ends of the bilateral common iliac veins (wherein the proximal end of the left common iliac artery is the distal end of the right common iliac artery crossing the left common iliac vein), and the proximal vessel of bilateral external iliac vein; the flow difference between the bilateral common iliac vein and the external iliac vein; and the pressure difference between the proximal end of the bilateral external iliac vein and the distal end of the inferior vena cava. The inferior vena cava plane was selected 2-3 cm away from the site where the bilateral common iliac converges into the inferior vena cava to reduce the impact of confluent blood flow on the measurement.

## 3. Data analysis

All statistical analyses were performed using SPSS version 23.0 statistical package (IBM, Armonk, NY, USA). Continuous variables are reported as mean ± standard deviation (SD) or median (quartile) and categorical variables as frequency or percentage of events. The bivariate Spearman’s test was used for correlation analysis. The relationships among the common iliac vein-inferior vena cava angle, stenosis rate, tortuosity, vessel area and iliac vein flow, velocity and pressure difference were analysed. A P value <0.05 was considered statistically significant.

## 4. Results

### 4.1 Volunteers’ characteristics

Thirty-three people were initially recruited to undergo 4D flow MRI scans, 3 of whom were excluded: 1 person failed the scan due to an excessively elevated heart rate (>110 beats/min), 1 person could not complete the scan due to large heart rate fluctuations, and 1 person could not fully display the bilateral iliac veins after reconstruction. A total of 30 people (60 limbs) were included in the study. The mean age of all volunteers was 24 years, the female to male ratio was 1:1, and the average BMI was 21.71±2.69 kg/m^2^(as shown in Table I).

**Table I.** Volunteer characteristics(n=30)

| Characteristic | Median (range) or % (No.) |
| --- | --- |
| Age, years | 24 (24~26) |
| Female gender | 15 (30) |
| Height, cm | $166.83 \pm 7.85$ |
| Weight, kg | $60.88 \pm 11.23$ |
| BMI, $\text{kg/m}^2$ | $21.71 \pm 2.69$ |

### 4.2 The median (interquartile range) of the Villalta score for all lower extremities was 0 (0-0), and the median venous clinical severity score (VCSS) was 0 (Figure 3)

**Figure 3:**
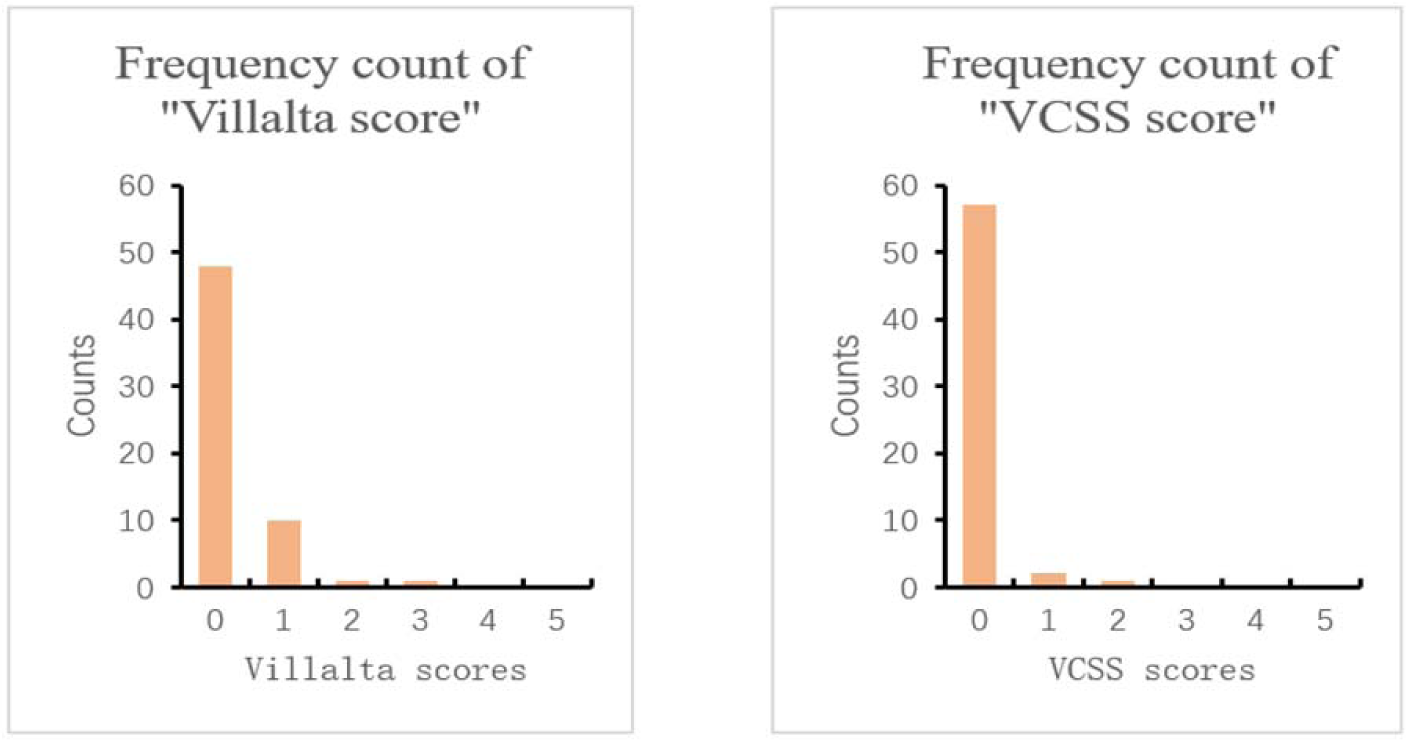
Histogram of frequency distribution of Villalta and VCSS scores for all sixty limbs.

### 4.3 Anatomical parameters

All limbs anatomical parameters are shown in Table II.

**Table II.**
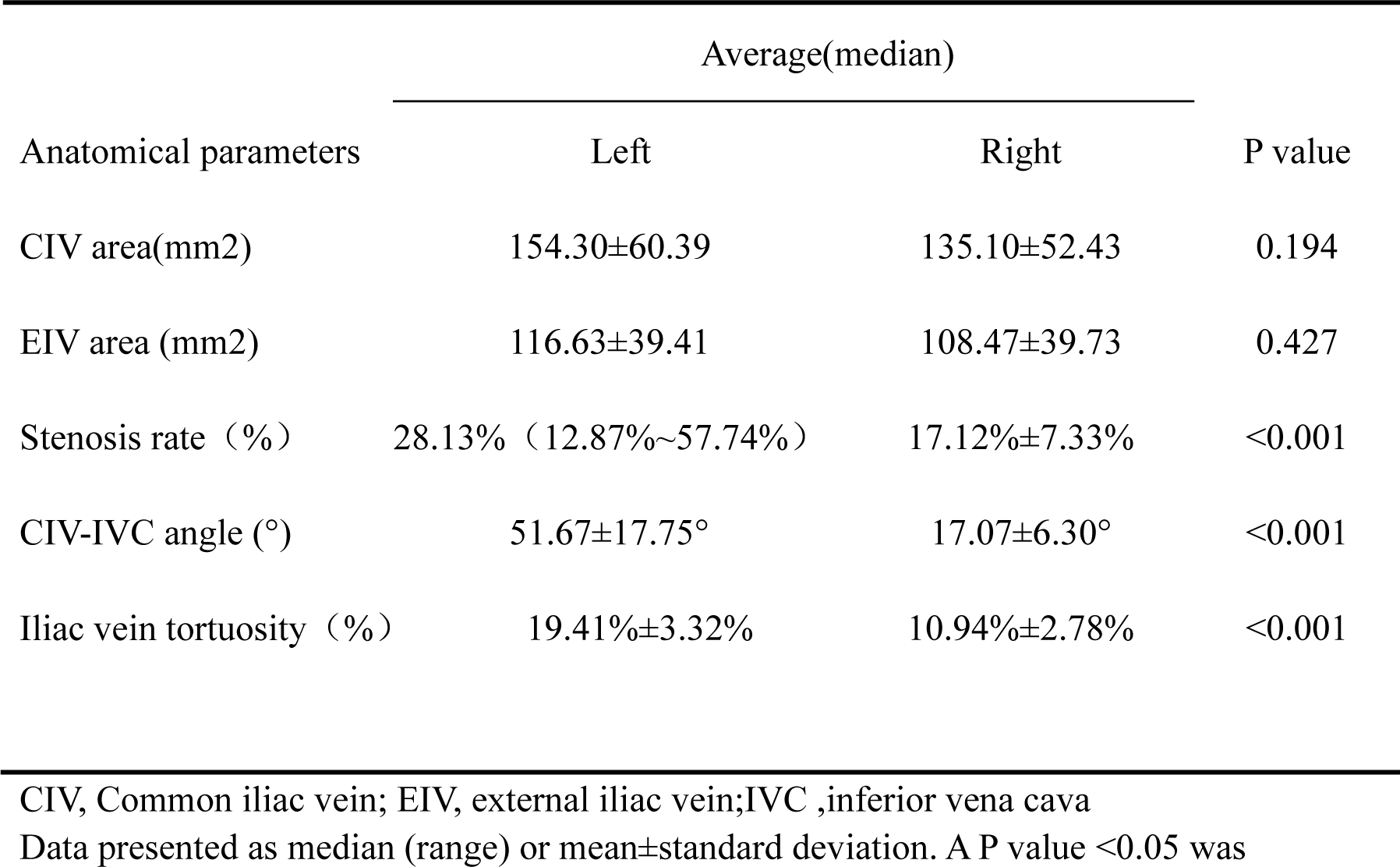
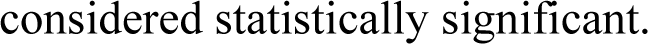
Anatomical parameters of all limbs(n=60)

| Anatomical parameters | Average(median) |  | P value |
| --- | --- | --- | --- |
|  | Left | Right |  |
| CIV area(mm2) | 154.30±60.39 | 135.10±52.43 | 0.194 |
| EIV area (mm2) | 116.63±39.41 | 108.47±39.73 | 0.427 |
| Stenosis rate (%) | 28.13% (12.87%~57.74%) | 17.12%±7.33% | <0.001 |
| CIV-IVC angle (°) | 51.67±17.75° | 17.07±6.30° | <0.001 |
| Iliac vein tortuosity (%) | 19.41%±3.32% | 10.94%±2.78% | <0.001 |

CIV, Common iliac vein; EIV, external iliac vein;IVC ,inferior vena cava
Data presented as median (range) or mean±standard deviation. A P value <0.05 was
considered statistically significant.

Vessel area: The median area of the inferior vena cava in all volunteers was 211.50 (170.00∼307.00) mm^2^, and the areas of the left common iliac and external iliac veins were larger than those of the opposite side, but the difference was not statistically significant.

Stenosis rate: The average stenosis rate of the left iliac vein was significantly higher than that of the right side. The median stenosis rate of all limbs was 23.35% (15.73%-30.29%); the average stenosis rate was 28.13%(12.87%∼57.74%) on the left side and 17.12%±7.33% on the right side (P<0.001).

Common iliac vein-inferior vena cava angle and tortuosity: The left iliac vein-inferior vena cava angle and tortuosity were greater than those of the right. The average iliac-vena cava angle of all limbs was 36.37±21.88°; on the left and right sides, the average angles were 51.67±17.75° and 17.07±6.30 (∼P<0.001), respectively. The median tortuosity of all limbs was 14.94% (10.07%∼19.70%); the average tortuosity of the left limb was 19.41%±3.32%, and that of the right limb was 10.94%±2.78%(∼P<0.001).

### 4.4 Haemodynamic parameters

All limbs haemodynamic parameters are shown in Table III.

**Talbe III.** Hemodynamic parameters of all limbs(n=60)

| Haemodynamic<br>parameters | Average(median) |  | P value |
| --- | --- | --- | --- |
|  | Left | Right |  |
| CIV blood flow ml/s | $6.70 \pm 2.88$ ml/s | $8.3 \pm 3.19$ ml/s | 0.045 |
| EIV blood flow ml/s | $5.08 \pm 1.85$ ml/s | $6.11 \pm 1.96$ ml/s | 0.041 |
| CIV-EIV flow<br>difference ml/s | $1.62 \pm 1.68$ ml/s | $2.19 \pm 2.69$ ml/s | 0.360 |
| CIV average flow<br>velocity cm/s | $9.27 \pm 3.43$ cm/s | $12.41 \pm 3.83$ cm/s | 0.001 |
| EIV average flow<br>velocity cm/s | $7.43 \pm 1.65$ cm/s | $9.94 \pm 2.34$ cm/s | <0.001 |
| EIV-IVC pressure<br>difference (mmHg) | $0.046 \pm 0.173$ mmHg | $-0.088 \pm 0.164$ mmHg | <0.001 |

CIV, Common iliac vein; EIV, external iliac vein; IVC, inferior vena cava

Flow velocity: We found that the average flow velocity of the left common iliac and external iliac veins was lower than that of the contralateral side. The average flow velocity of the inferior vena cava in all volunteers was 12.71±3.15 cm/s. The average flow velocity of the common iliac vein of all limbs was 10.84±3.94 cm/s, among which that of the left common iliac was 9.27±4.43 cm/s, and that of the right common iliac was 12.41±3.83 cm/s (P=0.001<0.01). The average flow velocity of external iliac vein of all limbs was 8.69±2.37 cm/s, among which the average velocity of the left iliac was 7.43±1.65 cm/s, and the average velocity of the right iliac was 9.94±2.34 cm/s (P<0.001).

Flow: The blood flow of the left common iliac and external iliac veins was lower than that of the opposite side. The mean blood flow of inferior vena cava of all volunteers was 14.53±4.74 ml/s. The mean blood flow of common iliac vein across all volunteers and that of the left and right common iliac veins were 7.50±3.12 ml/s, 6.70±2.88 ml/s, and 8.31±3.19 ml/s (P=0.045<0.05). For all limbs, the mean blood flow of external iliac vein was 5.60±1.96 ml/s, that of left external iliac was 5.08±1.85 ml/s, and that of right external iliac was 6.11±1.96 ml/s.

Flow difference: The average common iliac vein-external iliac vein flow difference of all limbs was 1.66 (0.675∼3.250 ml/s), while the flow differences between the left and right common iliac vein and the external iliac vein were 1.62±1.68 ml/s, and 2.19±2.69 ml/s(P=0.36>0.05), respectively. The right iliac vein flow difference was higher than that of the left iliac vein, but the difference was not statistically significant.

Pressure difference: We found that the left external iliac vein pressure was higher than that of the right side. The pressure difference between the proximal end of the bilateral external iliac vein and the proximal end of the inferior vena cava at rest was measured. The results showed that the average pressure difference on the left side was 0.046±0.173 mmHg, the average pressure difference on the right side was -0.088±0.164 mmHg(P=0.003<0.01), and the difference between the two sides (right vs left) was -0.13±0.22 mmHg.

### 4.5 The relationship between anatomy and haemodynamics

The relationship between anatomy and haemodynamics is shown in Table IV. We found correlations between pelvic vein anatomical parameters and haemodynamic parameters. A larger iliac vein area was correlated with a higher the blood flow; a higher iliac vein stenosis rate was correlated with a lower blood flow, leading to a decrease in the average external iliac vein velocity and an increase in the external iliac-inferior vena cava pressure difference; a greater common iliac vein-inferior vena cava angle was correlated with a lower iliac vein flow velocity and higher the external iliac vein-inferior vena cava pressure difference. A greater iliac vein tortuosity was correlated with a lower iliac vein velocity and higher external iliac vein-inferior vena cava pressure difference. Limbs with a greater tortuosity and angle had a lower common iliac vein flow, external iliac vein flow, average common iliac vein flow velocity, and average external iliac vein flow velocity and a higher external iliac-inferior vena cava pressure difference.

**Table IV.**
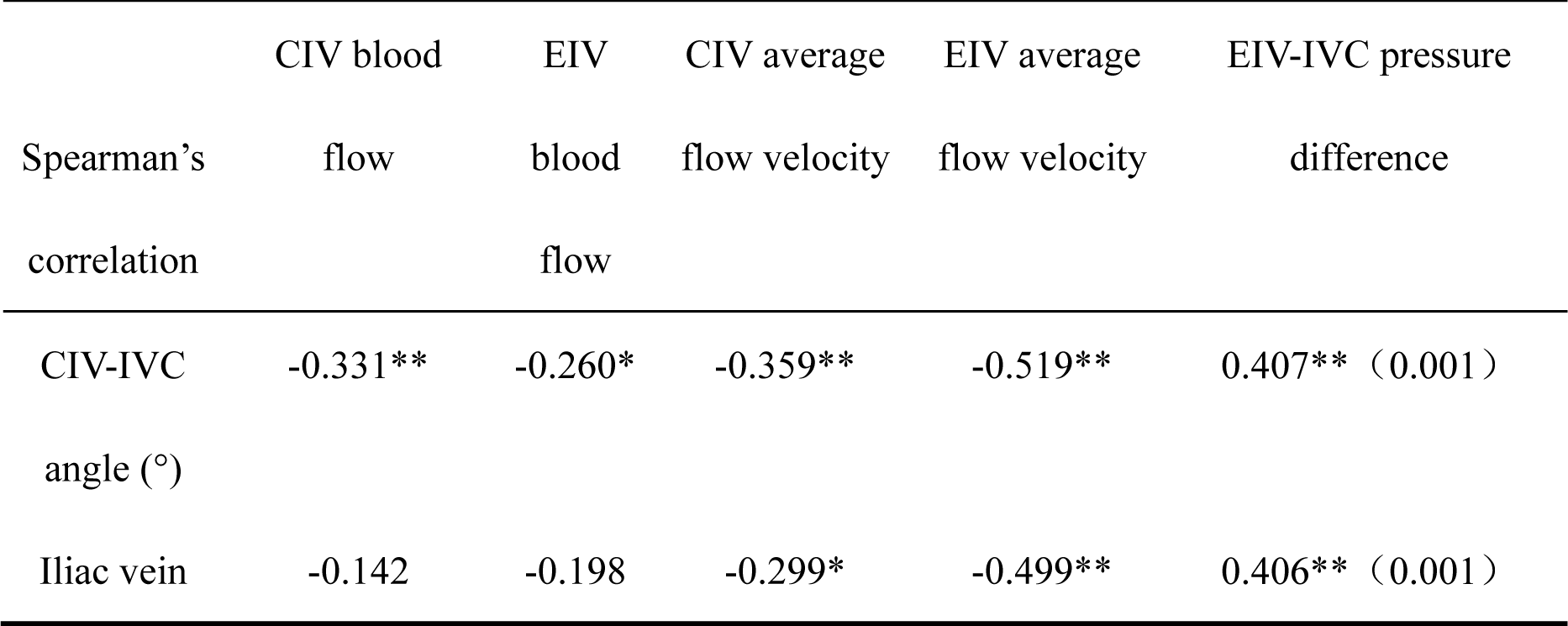
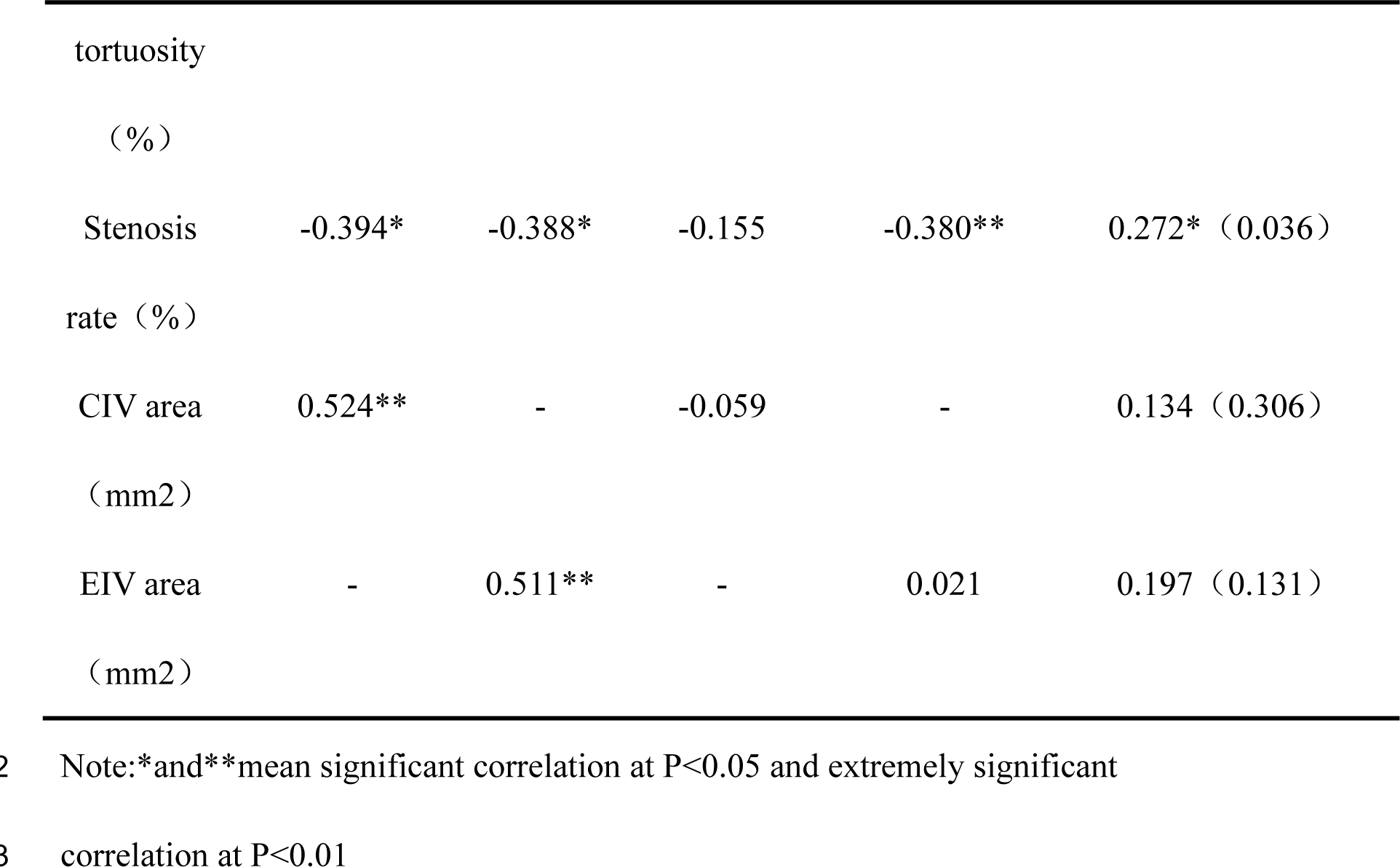
Correlation between anatomy and haemodynamics of iliac vein

|  | CIV blood | EIV | CIV average | EIV average | EIV-IVC pressure |
| --- | --- | --- | --- | --- | --- |
| Spearman's | flow | blood | flow velocity | flow velocity | difference |
| correlation |  | flow |  |  |  |
| CIV-IVC | -0.331** | -0.260* | -0.359** | -0.519** | 0.407** (0.001) |
| angle (°) |  |  |  |  |  |
| Iliac vein | -0.142 | -0.198 | -0.299* | -0.499** | 0.406** (0.001) |
| tortuosity |  |  |  |  |  |
| (%) |  |  |  |  |  |
| Stenosis | -0.394* | -0.388* | -0.155 | -0.380** | 0.272* (0.036) |
| rate (%) |  |  |  |  |  |
| CIV area | 0.524** | - | -0.059 | - | 0.134 (0.306) |
| (mm<sup>2</sup>) |  |  |  |  |  |
| EIV area | - | 0.511** | - | 0.021 | 0.197 (0.131) |
| (mm<sup>2</sup>) |  |  |  |  |  |

Note:\*and\*\*mean significant correlation at P<0.05 and extremely significant
correlation at P<0.01

## 5. Discussion

Venous stasis in the lower extremities is considered to be an important haemodynamic change in the occurrence and development of CVD. Some studies have reported that the obstruction and reflux disorder of the lower extremities caused by iliac vein stenosis are a reason for venous insufficiency of the lower extremities and the subsequent formation of CVD^2, 3^. The clinical use of iliac vein stents to correct stenosis has been shown to be effective^4–6^, but studies have shown that iliac vein stenosis rates do not exactly match symptom severity^7^. Asymptomatic iliac vein stenosis is a normal anatomical variation rather than a pathological change^8, 9^.

In recent years, studies have found that abnormal haemodynamics are closely related to the occurrence and development of vascular diseases. The common iliac and external iliac veins are the channels for venous reflux of the lower extremities. We believe that the anatomical parameters of the iliac veins, such as vessel area, stenosis rate, and vessel course, may affect iliac vein haemodynamic parameters, such as flow, velocity, and pressure, which will lead to progression in CVD. In addition, in clinical practice, the prevalence of CVD on the left and right sides has been shown to be inconsistent^10, 11^, and we hypothesize that this inconsistency is related to a certain degree with the anatomical differences between the bilateral limbs.

At present, the diagnosis and evaluation of vascular diseases are very dependent on imaging data, such as Doppler ultrasound, computed tomography angiography (CTA), magnetic resonance angiography (MRA), digital subtraction angiography (DSA), and intravascular ultrasound (IVUS), but few of the above methods can accurately, objectively and dynamically collect the haemodynamic parameters of the patients. 4D flow MRI has great application prospects in the research of vascular diseases due to its noninvasiveness, lack of radiation, lack of contrast agent enhancement, and wide coverage. It can provide a large number of haemodynamic parameters and visualize blood flow status and changes and allows analysis of postprocessed images offline for retrospectively assessing blood flow parameters^12^. The ability of 4D flow MRI to perform qualitative and quantitative analysis allows a comprehensive assessment of pelvic venous blood flow and can help clarify the relationship between haemodynamic changes and the development of CVD. In this paper, 4D flow MRI was first used to collect the anatomical and haemodynamic parameters of the pelvic vein and to explore the anatomical and haemodynamic characteristics of the iliac vein in asymptomatic people and their correlations, providing a reference for further exploring abnormal changes in fluid dynamics.

We measured the anatomical parameters of the iliac vein in asymptomatic individuals and found that the average angle of the common iliac vein as it converged into the inferior vena cava in all limbs was 36.37±21.88°, with average angles on the left and right of 51.67±17.75° and 17.07±6.30° (P<0.001), respectively. The angle at which the left common iliac vein converged into the inferior vena cava was larger than that on the right side. This suggests that it may be more appropriate to choose about 50° for the bevelled opening angle of the iliac vein-specific stent in current clinical practice. The median (interquartile range) tortuosity of all limbs was 14.94% (10.07%-19.70%), among which the mean tortuosity of the left iliac vein was 19.41%±3.32%, and the mean tortuosity of the right iliac vein was 10.94%±2.78%(P<0.001), showing that the tortuosity of the left iliac vein was greater than that of the contralateral side. The course of the right common iliac vessels is steeper and straighter than the left, which is consistent with the research results of Arjun Jayaraj^13^ et al. The tortuosity of the left iliac vein is large, a greater tortuosity requires higher compliance for the middle section of the iliac vein stent, as this would allow better adaptation to the tortuosity of the iliac vein and adherence to the vein wall, suggesting that support and flexibility should be considered when selecting materials for the design of iliac vein stents. Raju^14^ et al. used colour Doppler ultrasound to measure the common iliac and external iliac diameters of healthy volunteers, obtaining values of 13.3 (11-15.8) mm and 11.5 (8.7-13.9) mm, respectively; this study found that the average diameters of left and right common iliac veins of the volunteers were 14.02 mm and 12.19 mm, respectively, while the average diameters of the left and right external iliac veins were 13.12 mm and 11.75 mm, respectively, which may also serve as reference values for the design of iliac vein stents.

Iliac vein stenosis is very common worldwide, but most people have iliac vein stenosis without obvious clinical symptoms. Iliac vein compression syndrome (IVCS) often occurs in the left lower extremity and is more common in women than in men^15^. Kibbe et al.^8^ found that among asymptomatic patients with lower extremity compression, approximately 24% had left common iliac vein compression greater than 50%. In this study, the iliac vein stenosis rate was >50% in 20% (6/30) of the left limbs of all volunteers as measured by 4D flow MRI, consistent with the findings of Kibbe et al. The mean stenosis rate of the left limb was higher than that of the right side (34.04%±13.74% vs. 17.12%±7.33%, P<0.001).

Currently, the relationship between iliac vein anatomy and haemodynamics is unclear. We used 4D flow MRI to measure the mean velocity and blood flow of the bilateral iliac veins and the pressure difference between the external iliac vein and inferior vena cava in an asymptomatic population. This study found significant correlations between iliac vein anatomy and haemodynamic parameters. A larger iliac vein area was correlated with a higher blood flow; a larger common iliac vein-inferior vena cava angle was correlated with a lower iliac vein velocity and a higher external iliac-inferior cavity pressure difference. Finally, a greater iliac vein tortuosity was correlated with a lower iliac vein velocity and a higher external iliac vein-inferior vena cava pressure difference.

This study found that an iliac vein stenosis rate of more than 50% affected the blood flow and pressure differences of the iliac vein. Specifically, compared with a stenosis rate of less than 50%, the absolute blood flow of the iliac vein decreased. While the relative blood flow (the difference between the common iliac and external iliac blood flows) decreased, the venous pressure increased, and the average iliac vein velocity decreased. However, none of these differences was significant, suggesting that compensatory velocity acceleration may occur at the site of venous stenosis (Table V). According to Poiseuille’s law, changes in intravascular flow velocity and pressure differences will lead to changes in blood flow. The flow velocity can be compensatorily increased at the site of stenosis and decreased at sites near the stenosis, so the flow velocity does not objectively reflect the status of venous reflux. Another study by our center^16^ suggested that changes in the common iliac-external iliac flow difference at the lower extremities, that is, a decrease in relative flow, may be related to the severity of the patient’s symptoms. A low or even negative flow difference may be associated with more severe limb symptoms (higher Villalta scores).

**Table V.**
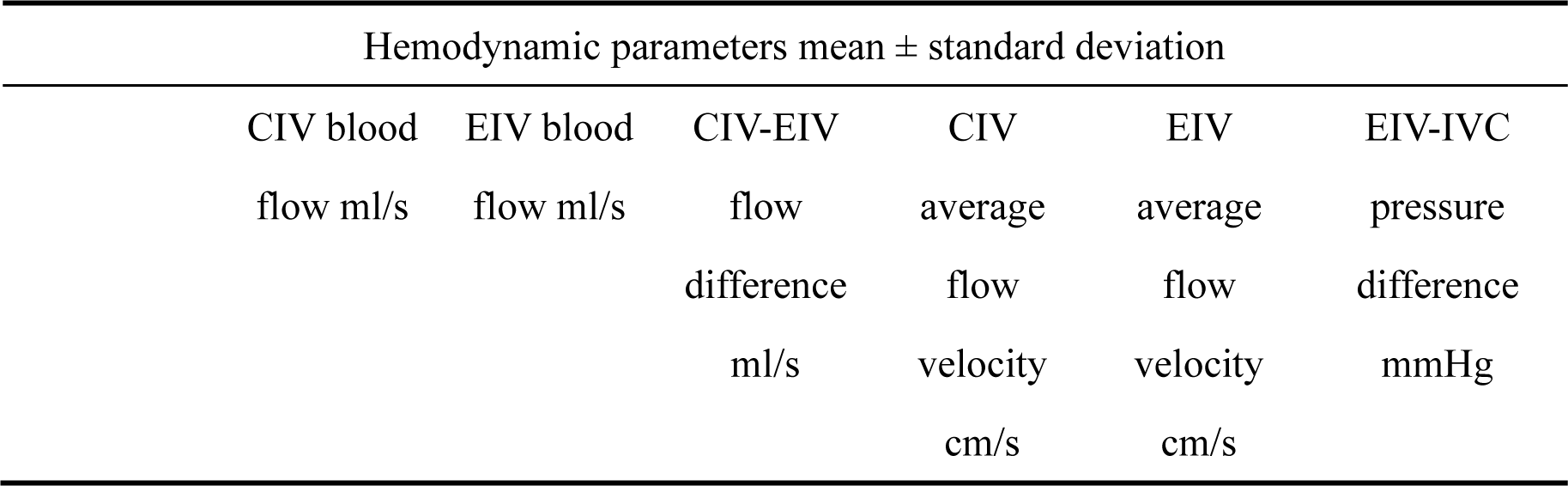
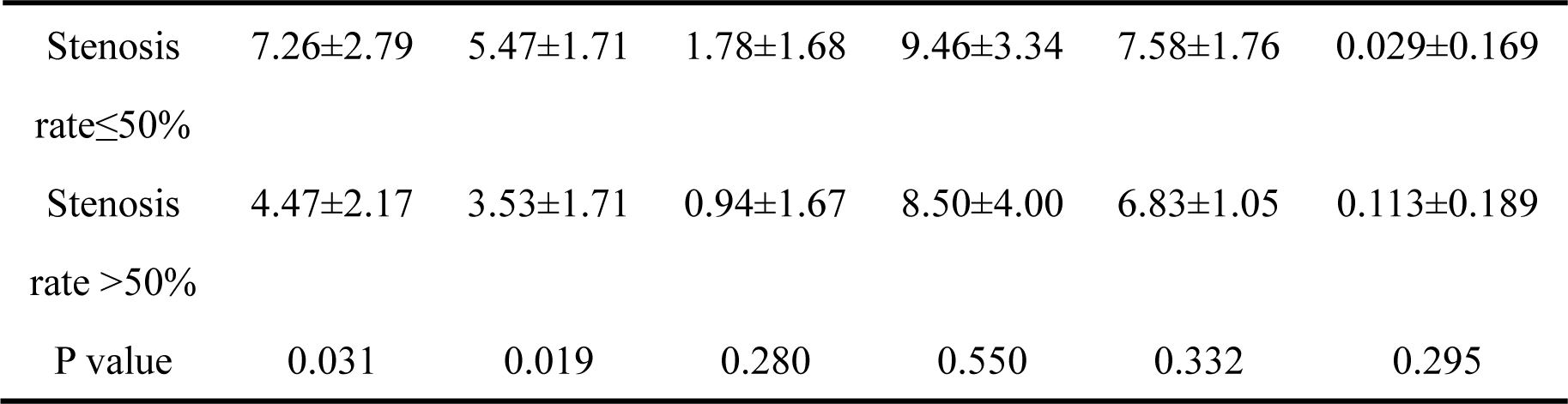
Correlation of degree of stenosis and hemodynamics

Twenty percent of the volunteers had a left iliac vein stenosis rate of >50% but no symptoms related to lower extremity venous insufficiency (such as varicose veins or oedema). We believe that stenosis can lead to changes in iliac vein haemodynamics, but the reason for the lack of clinical symptoms is related to the fact that although a certain degree of iliac vein stenosis is a physiological anatomical structure, it is also a potential pathological change. The changes in venous flow due to venous stenosis in the normal population may have a “threshold effect”, and a decrease in venous flow may not be sufficient to cause symptoms or signs related to the veins of the lower extremities. Symptoms associated with venous insufficiency of the lower extremity may be induced if the stenosis causes the flow or flow difference to reach this threshold.

After stratified analysis of iliac vein angle and tortuosity, we found that the common iliac vein flow, external iliac vein flow, average common iliac vein velocity, and average external iliac vein flow velocity were all lower in limbs with greater iliac vein angles and tortuosity values, while the external iliac-inferior vena cava pressure difference was higher (Table VI, VII). Compared with those of the right side, the tortuosity and common iliac vein-inferior vena cava angle of the left side were larger. The average flow and velocity in the right common and external iliac veins were higher than in the left limb, and the pressure in the left external iliac vein was higher than that in the right side. The average pressure difference between the left external iliac vein and the right external iliac vein was 0.13±0.22 mmHg. We believe that the differences in the bilateral iliac vein tortuosity and venous inflow angle may also be a reason for the difference in haemodynamic parameters of bilateral iliac veins, which can explain why lower extremity venous insufficiency in CVD patients mostly occurs in the left side. Lower extremity venous insufficiency was also more common on the left side in patients with no iliac vein stenosis on angiography. This is closely related to the larger angle of the left common iliac vein as it converges into the inferior vena cava and its larger tortuosity.

**Table VI.**
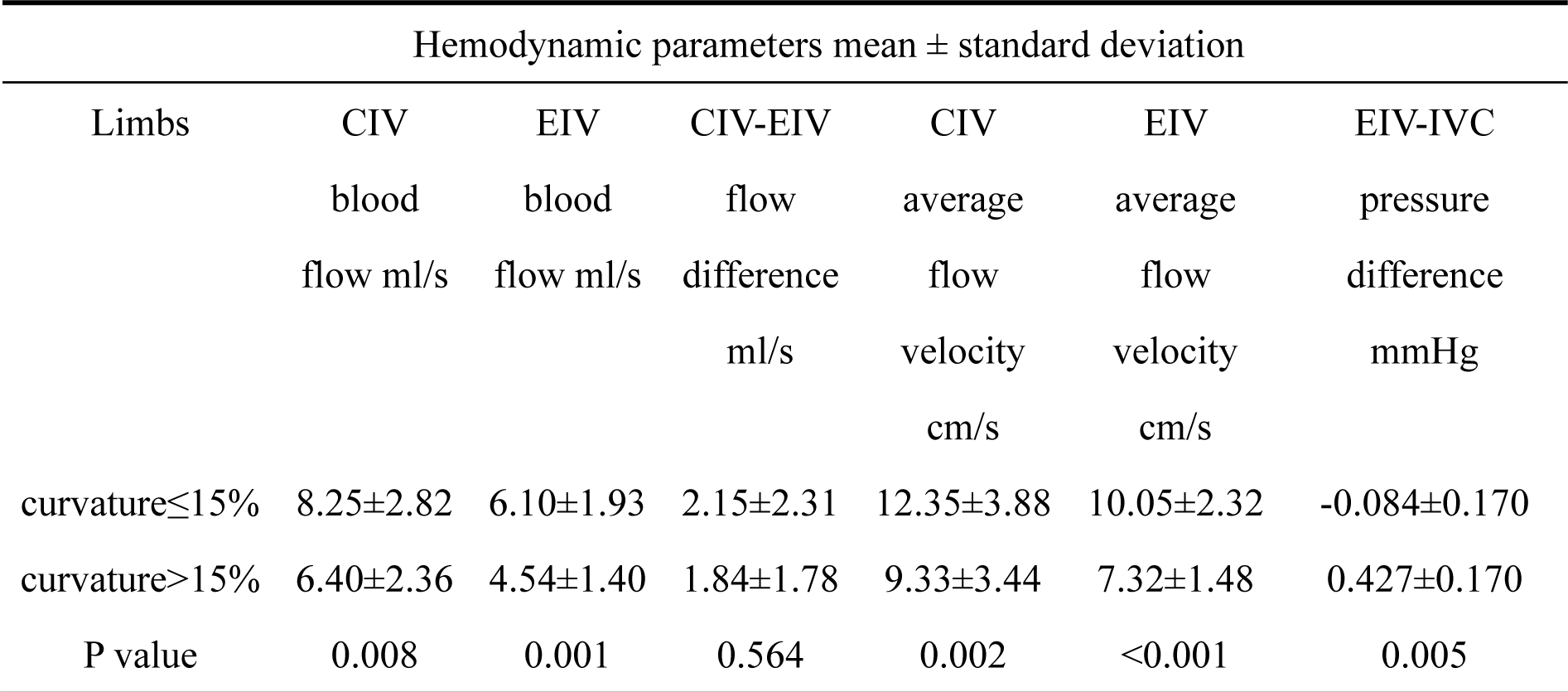
Correlation of curvature and hemodynamics

| Hemodynamic parameters mean ± standard deviation | | | | | | |
| --- | --- | --- | --- | --- | --- | --- |
| Limbs | CIV<br>blood<br>flow ml/s | EIV<br>blood<br>flow ml/s | CIV-EIV<br>flow<br>difference<br>ml/s | CIV<br>average<br>flow<br>velocity<br>cm/s | EIV<br>average<br>flow<br>velocity<br>cm/s | EIV-IVC<br>pressure<br>difference<br>mmHg |
| curvature≤15% | 8.25±2.82 | 6.10±1.93 | 2.15±2.31 | 12.35±3.88 | 10.05±2.32 | -0.084±0.170 |
| curvature$>$15% | 6.40±2.36 | 4.54±1.40 | 1.84±1.78 | 9.33±3.44 | 7.32±1.48 | 0.427±0.170 |
| P value | 0.008 | 0.001 | 0.564 | 0.002 | $<0.001$ | 0.005 |

**Table VII.** Correlation of angle of common iliac vein-inferior vena cava and hemodynamics

| Hemodynamic parameters mean ± standard deviation | | | | | | |
| --- | --- | --- | --- | --- | --- | --- |
| Limbs | CIV blood<br>flow ml/s | EIV blood<br>flow ml/s | CIV-EIV<br>flow<br>difference | CIV<br>average<br>flow | EIV<br>average<br>flow | EIV-IVC<br>pressure<br>difference |

|  |  |  | ml/s | velocity<br>cm/s | velocity<br>cm/s | mmHg |
| --- | --- | --- | --- | --- | --- | --- |
| Angle≤37° | 8.10±2.67 | 5.88±1.85 | 2.20± | 12.01± | 9.48±2.34 | -0.092±0.15 |
|  |  |  |  | 3.67 |  |  |
| Angle>37° | 6.08±2.42 | 4.43±1.48 | 1.68± | 8.96± | 7.41±1.85 | 0.093±0.16 |
|  |  |  |  | 3.68 |  |  |
| P value | 0.005 | 0.002 | 0.166 | 0.003 | 0.001 | 0.000 |

## 6. Conclusion

The anatomical structure of the pelvic vein, including the iliac vein area, common iliac vein-inferior vena cava angle, iliac vein tortuosity, and iliac vein stenosis rate, are important factors that affect the haemodynamic changes in the pelvic vein. There are correlations between parameters related to anatomical structure and haemodynamic changes in the pelvic veins. Differences in the anatomy and haemodynamics of the bilateral pelvic veins may be one of the reasons why the left extremity is more prone to CVD-related symptoms.

## 7. Limitations

It is important to bear in mind the limitations of this study. First, the sample size of this study was small. Second, our study failed to explain the effect of circadian rhythm, respiratory rate, body position, and exercise on blood flow. In addition, when the participant is in the supine position, the 4D flow MRI scan is not dynamic and may not reflect the anatomic structures when the participant is standing and moving. We also did not acquire measurement data for IVCS patients and older persons as controls. More prospective studies with high quality and larger sample sizes should be designed in the future.

## Data Availability

The data that support the findings of this study are available on request from the corresponding author?upon reasonable request.

